# The Cholinergic Anti-Inflammatory Reflex as a Function of Depression Severity in Adolescent Non-Suicidal Self-Injury

**DOI:** 10.1101/2025.01.02.25319936

**Authors:** Thomas P. Nguyen, Luise Baumeister-Lingens, Anna Markser, Anna-Sophia Rösch, Michael Kaess, Julian Koenig

## Abstract

A growing body of research has demonstrated a link between non-suicidal self-injury (NSSI) and mood disorders with pro-inflammatory states. The cholinergic anti-inflammatory reflex is a neural pathway that helps modulate the body’s inflammatory response and has been posited as a potential avenue for adjuvant treatment in mental disorders. However, no study to date has investigated the functioning of the cholinergic anti-inflammatory reflex in a clinical adolescent population with NSSI. This study measured heart rate variability (HRV; a proxy for vagus nerve activity), inflammatory markers (leukocytes, c-reactive protein (CRP), interleukin-6 (IL- 6)) and several clinical measures in female adolescents with NSSI (n = 154) along with healthy controls (n = 46). Statistical analyses tested for group differences and correlations between HRV, inflammatory markers and depression in the NSSI group. Mediation analyses were conducted to test direct and indirect effects. The NSSI group showed greater depressive symptoms and leukocyte levels, but lower HRV compared to the control group. In the full sample, depression severity was positively correlated with leukocyte and CRP levels and negatively correlated with HRV. HRV was also negatively correlated with leukocyte and CRP levels. Depression severity mediated the association between leukocytes and HRV. Overall, this study supports the notion that lower vagal activity is associated with increased inflammatory markers in a sample of adolescents with NSSI which suggests altered functioning of the cholinergic anti-inflammatory reflex.

## Introduction

Non-suicidal self-injury (NSSI) refers to the deliberate and self-inflicted damage of body tissue, where there is no suicidal intent nor social and cultural sanctioning (Kaess et al., 2021a). The prevalence of a single episode of NSSI in adolescents in non-clinical samples is 17.2%, which is higher than in younger and older adults (Swannell et al., 2014). In clinical adolescent samples, NSSI has been found to predict future suicide attempts more strongly than previous suicide attempts (Asarnow et al., 2011; Wilkinson et al., 2011). NSSI has also been consistently associated with both depressive symptoms (Marshall et al., 2013) and a diagnosis of depression (Ghinea et al., 2020) though the directionality of this association remains unclear.

Neurobiological studies investigating the relationship between the autonomic nervous system (ANS) and NSSI suggest that the sympathetic nervous system may be more dominant in individuals with NSSI (Kaess et al., 2021a; Koenig et al., 2023). Heart rate variability (HRV), which refers to the natural beat-to-beat variability in heart rate (HR), indexes cardiac parasympathetic activity within the ANS. A higher HRV indicates greater flexibility of the ANS to adapt to changes in the environment (Appelhans & Luecken, 2006). Meanwhile, a lower HRV is considered to be a transdiagnostic marker for several mental illnesses (e.g., depression, anxiety disorders, bipolar disorder and schizophrenia) due to the hyperarousal of the sympathetic nervous system which leads to deficits in cognitive control and self-regulation (Baumeister-Lingens et al., 2023; Beauchaine & Thayer, 2015; Paniccia et al., 2017).

Depression has also been associated with a lower HRV. In a sample of adolescents with NSSI, one study found that resting state HRV was inversely correlated with BPD symptomatology (Koenig, et al., 2017). Further analyses of this sample also revealed that this relationship was maintained longitudinally (Koenig et al., 2018). These findings which suggest an inverse relationship between BPD symptoms and HRV have also been independently replicated (Hedinger et al., 2023). With respect to depression, clinically depressed children and adolescents have been shown to have significantly lower levels of HRV when compared to healthy controls (Koenig et al., 2016). Reductions in HRV have also been found to precede the development of depressive symptomatology in adults (Jandackova et al., 2016), suggesting that autonomic dysfunction may be related to the pathophysiology of depression. With respect to sex, meta-analytic data in healthy samples suggests that girls display lower vagally-mediated HRV at resting-state than boys due to hormonal changes during puberty and sex-differences in physical activity levels (Koenig, Rash, et al., 2017), potentially underlying the increased risk for depression in girls.

Research has also linked the immune system with the pathophysiology of depression. A recent meta-analysis reported that the relationship between depression and inflammation may be bidirectional in children and adolescents (Colasanto et al., 2020). The meta-analysis also reported significant associations between the inflammatory markers c-reactive protein (CRP) and interleukin-6 (IL-6) and concurrent depression in cross-sectional studies of children and adolescents (Colasanto et al., 2020). IL-6 is produced early in the inflammatory process and triggers CRP which is involved in cytokine production and regulation of the complement system (Lamers et al., 2019). These inflammatory markers are thought to promote sickness behaviours (e.g., fatigue, anorexia, anhedonia) and induce brain alterations such as the disruption of synaptic plasticity and the activation of corticotropin-releasing hormone (Lamers et al., 2019; Maes et al., 2012). Moreover, studies have also found significantly higher leukocyte levels in adolescents with depression compared to healthy controls (Puangsri & Ninla-Aesong, 2021; Uçar et al., 2018). Earlier studies investigating the relationship between NSSI and inflammation failed to replicate the same associations with CRP (Kindler et al., 2022) and IL-6 (Kim et al., 2020), although the results were mixed with respect to leukocytes (Kindler et al., 2022; Zheng et al., 2022). However, other inflammatory markers (e.g., tumour necrosis factor-α, monocyte-lymphocyte ratio) have been found to be elevated in adolescents with NSSI (Kim et al., 2020; Zheng et al., 2022).

Although the role of the nervous and immune systems in depression has been extensively researched, the interplay between the two systems has not yet been sufficiently investigated in clinical samples. It is thought that the ANS may regulate the body’s inflammatory response through a cholinergic anti-inflammatory pathway called the ‘inflammatory reflex’ (Tracey, 2002). Following an inflammatory response, afferent signals are sent to the nucleus tractus solitarius via the vagus nerve (Tracey, 2002). The efferent signal from the vagus nerve releases acetylcholine which inhibits the synthesis of pro-inflammatory cytokines (Tracey, 2002). Therefore, dysregulation of the ANS, as indexed by low HRV, would lead to an excess production of pro-inflammatory cytokines. A recent meta-analysis of human studies revealed a negative association between HRV and inflammatory markers, with CRP and leukocytes demonstrating the strongest associations, while IL-6 also also showed significant negative associations (Williams et al., 2019). While these findings support the concept of the ‘inflammatory reflex’, no study to date has investigated these mechanisms in adolescents who engage in NSSI.

While the relationship between the nervous and immune systems in the pathophysiology of depression has received much attention in recent years, little research to date has sought to investigate the link between both systems, despite knowledge that there may be an underlying, synergistic process. Given the high prevalence of depression and single episodes of NSSI in adolescence, it is important to understand which biomarkers predict these disorders and whether the ‘inflammatory reflex’ is implicated in these populations. Our study aimed to investigate the differences and relationships between resting-state HRV (as a proxy of vagal nerve activity), inflammatory markers (leukocytes, IL-6 and CRP) and depressive symptoms in adolescent girls engaging in NSSI compared to a control sample of healthy adolescent girls.

We firstly hypothesized that adolescents engaging in NSSI would exhibit greater depressive symptoms, increased inflammatory markers and lower HRV compared to the control group. Secondly, we hypothesized that on a continuum, depression severity would be positively associated with inflammatory markers and negatively associated with HRV. Finally, we sought to investigate whether reductions in HRV were associated with increases in inflammatory markers, in line with the ‘inflammatory reflex’ and whether depression severity mediates this association.

## Methods

### Participants, Eligibility Criteria and Procedures

In this cross-sectional, case-control study, patients engaging in NSSI were drawn from AtR!Sk (*Ambulanz für Risikoverhalten und Selbstschädigung*), a specialized German outpatient clinic for risk-taking and self-harm behaviour at the University Hospital Heidelberg, Germany (Kaess et al., 2017). Following an initial assessment at the clinic, patients were invited to participate in AtR!Sk-Bio, a nested cohort study designed to identify biological correlates of NSSI in adolescence. The scientific evaluation of AtR!Sk was approved by Faculty of Medicine Ethics Committee at the University of Heidelberg (IRB approval number S-449/2013, and the additional neurobiological assessments: IRB approval number S-514/2015).

Patients who engaged in NSSI initially underwent a structured psychiatric diagnostic interview with a trained psychologist. They were then given the option of participating in the present study. Patients were invited to participate in the study if they had completed this diagnostic assessment, displayed risk-taking behaviours, were between the ages of 12-17 years and had themselves and their caregivers provide written informed consent. Patients were excluded if they exhibited acute psychotic symptoms or had insufficient speech comprehension. For the present analyses, patients were only included if they had engaged in at least five incidents of NSSI in the past twelve months, as defined in the DSM-5 (American Psychiatric Association, 2013).

Recruitment for healthy controls occurred through public advertisements. Potential participants were then initially assessed over telephone for suitability for inclusion into the study. Following an explanation of the study, a letter was sent out including further information about the study, written consent forms and self-conducted baseline questionnaires. Control participants also undertook an adapted version of the diagnostic assessment to the AtR!Sk patient group. Participants were eligible for the study if they had no history of NSSI, no endorsement of any mental disorder with concurrent treatments prior to participating in the study and had themselves and their caregivers provide written informed consent.

Once all participants from both groups had provided written informed consent and completed their initial clinical diagnostic assessment, they attended a second appointment to complete all neurobiological assessments. Second appointments were scheduled at 0800 am to ensure fasting blood draws and consistency with circadian rhythms. All participants had their height and weight measured to determine body mass index (BMI) and answered questions regarding their drug, alcohol and smoking status, recent medical history (in the last 3 months), regular medications and fasting status to account for any variations with the blood draw. All participants received 40€ at the conclusion of the biological assessments.

### Clinical Measures

During the first study site visit, the Mini International Neuropsychiatric Interview for Children and Adolescents (MINI-KID) was conducted to assess psychiatric diagnoses. The MINI-KID is a reliable and valid semi-structured diagnostic interview that was used to diagnose common DSM-IV axis I psychiatric disorders in the present study (Sheehan et al., 2010). The frequency and severity of NSSI was measured using the German version of the Self-Injurious Thoughts and Behaviours Interview (SITBI-G) which is comparable to the original version and demonstrates good psychometric properties (Fischer et al., 2014). Self-reported depressive symptoms were assessed through the Depression Inventory for Children and Adolescents (DIKJ) (Stiensmeier-Pelster et al., 2000). This inventory demonstrates excellent psychometric properties and contains 26 items which are based off the DSM-IV criteria for depression (Stiensmeier-Pelster et al., 2000).

### Blood Draws

Fasting blood samples for CRP (mg/l), leukocytes (/nl) and IL-6 (pg/ml) were collected in the morning of the second appointment by venepuncture. They were then sent to the central laboratories of the Heidelberg University Hospital for further analyses. CRP concentrations were examined by immunoassays from a lithium heparin tube. Leukocyte levels were assessed through the quantitative measure of scattered light in a continuous flow from an EDTA tube. IL-6 levels were determined through a chemiluminescence immunoassay from a blood serum tube. For CRP (86,7%) and IL-6 (80,3%), the majority of values were below the detection threshold. Thus, results are reported for all available data and values above the detection threshold respectively.

### Heart Rate and Heart Rate Variability

The reporting of HRV processing and analyses in this study adheres to the Guidelines for Reporting Articles on Psychiatry and Heart rate variability (Quintana et al., 2016). Electrocardiogram (ECG) recordings were taken using the Movisens ECG-Move Sensor (Movisens, GmbH, Karlsruhe) during a 5-min baseline while participants completed a *Color Detection Task* (CDT; Jennings, Kamarck, Stewart, Eddy, & Johnson, 1992). Patients wore an elastic chest belt with a sensor attached to the strap and integrated electrodes. During the ECG recording, participants were comfortably seated and were instructed to uncross their legs and breathe normally. ECG signals were recorded at a sampling rate of 1024 *Hz*. Data were visually inspected after every recording using the unisens viewer (version: 2.0) and saved in the csv format. ECG data were further processed in Kubios HRV 3.0 Premium (Tarvainen et al., 2014). R-Peak detection was manually corrected and artifacts were removed. Smoothing priors were selected as detrending method (λ 500) for IBI data. Kubios output was saved in the txt format for later automated readout of corrected inter-beat-intervals (IBIs) and analysis of heart rate variability (HRV) in R (Martínez et al., 2017). IBIs corresponding to a mean HR < 30 or > 200 bpm were discarded and data were segmented in accordance with experimental conditions. The square root of the mean squared difference of successive IBIs (RMSSD) measured in *ms*, a time-domain measure of HRV indexing vagal activity (Task Force of the European Society of Cardiology and the North American Society of Pacing and Electrophysiology, 1996) and the mean heart rate (HR), were calculated.

### Statistical analyses

Pre-processing of data as well as statistical analyses were performed with Stata/SE (version 17.0; StataCorp LP, College Station, TX, US), if not further specified. First, descriptive statistics were calculated for demographic data, psychopathological variables and physiological indices of interest. To determine differences between the two study groups, we performed t-tests of group differences in the variables HR and HRV, inflammatory markers CRP, leukocytes and IL-6, as well as depression severity. To identify potential covariates to include in subsequent analyses investigating the relationships between cardiac vagal control (HRV), inflammatory state and depression in adolescents engaging in NSSI, we calculated pairwise correlations for HRV and HR, inflammatory markers CRP, leukocytes, IL-6, depression severity, number of BPD symptoms and frequency of NSSI within the past year.

Using SPSS (ver. 29.0.1.0, IBM, Chicago, IL, USA), we employed the PROCESS custom dialog (Hayes, 2012) to investigate the independent mediating role of depression severity in the relationship between resting-state HRV and various inflammatory markers. Within the ’Model 4’ framework of PROCESS, we defined resting-state HRV as the independent variable (IV), depression severity as mediating variable (M). We performed three mediation models, each with one of the three inflammatory markers as the dependent variable (DV). To assess the significance of the mediating or indirect effect, bootstrapping confidence intervals (CI) with a 95% interval and a sampling rate of 5000 (Preacher et al., 2007; MacKinnon et al., 2004 for bootstrapping details) were used. The reported statistics include unstandardized betas (B), standard errors, and the bootstrapping CI (lower limit, upper limit) for each path in the model. Non-zero-inclusive CI values were considered indicative of statistical significance. All tests were two-tailed, with a predetermined significance level of α = 0.05.

## Results

A total of n = 154 female adolescents with NSSI and n = 46 female healthy controls were included in the final analyses. There were no statistically significant differences on age or BMI between groups. However, the healthy control group were less likely to smoke and be on prescribed medications and were more likely to attend the highest track of secondary school. The NSSI group fulfilled more BPD criteria than the healthy control group and reported significantly higher depression severity, as well as leukocyte levels. Conversely, the healthy controls had significantly higher HRV (*Figure 1A*). The group differences for leukocyte levels (*Figure 2A*), depression and HRV remained statistically significant on mixed-effects multilevel regression when controlling for group differences in smoking and medication use (*Supplementary Table 1*).

**Figure 1:**
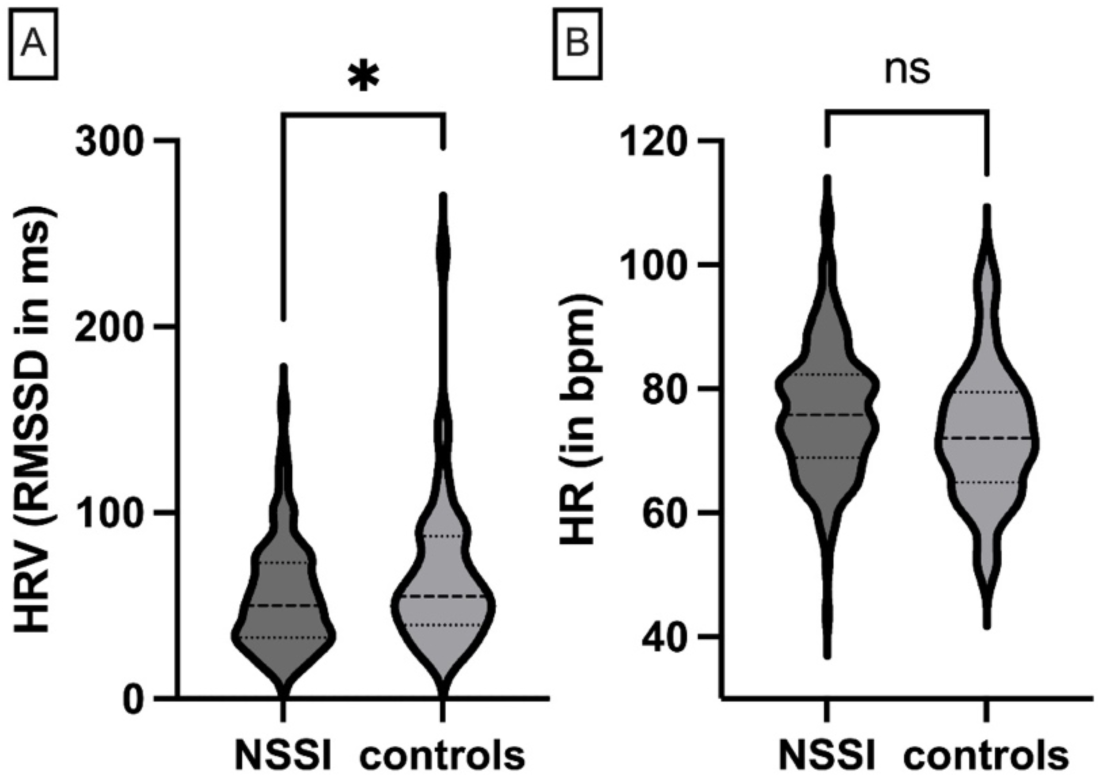
Hear Rate and its Variability by Group; HRV: heart rate variability; RMSSD: root of the mean squared difference of successive inter-beat intervals in milliseconds; HR: heart rate; NSSI: non-suicidal self-injury; ns: non-significant

**Figure 2:**
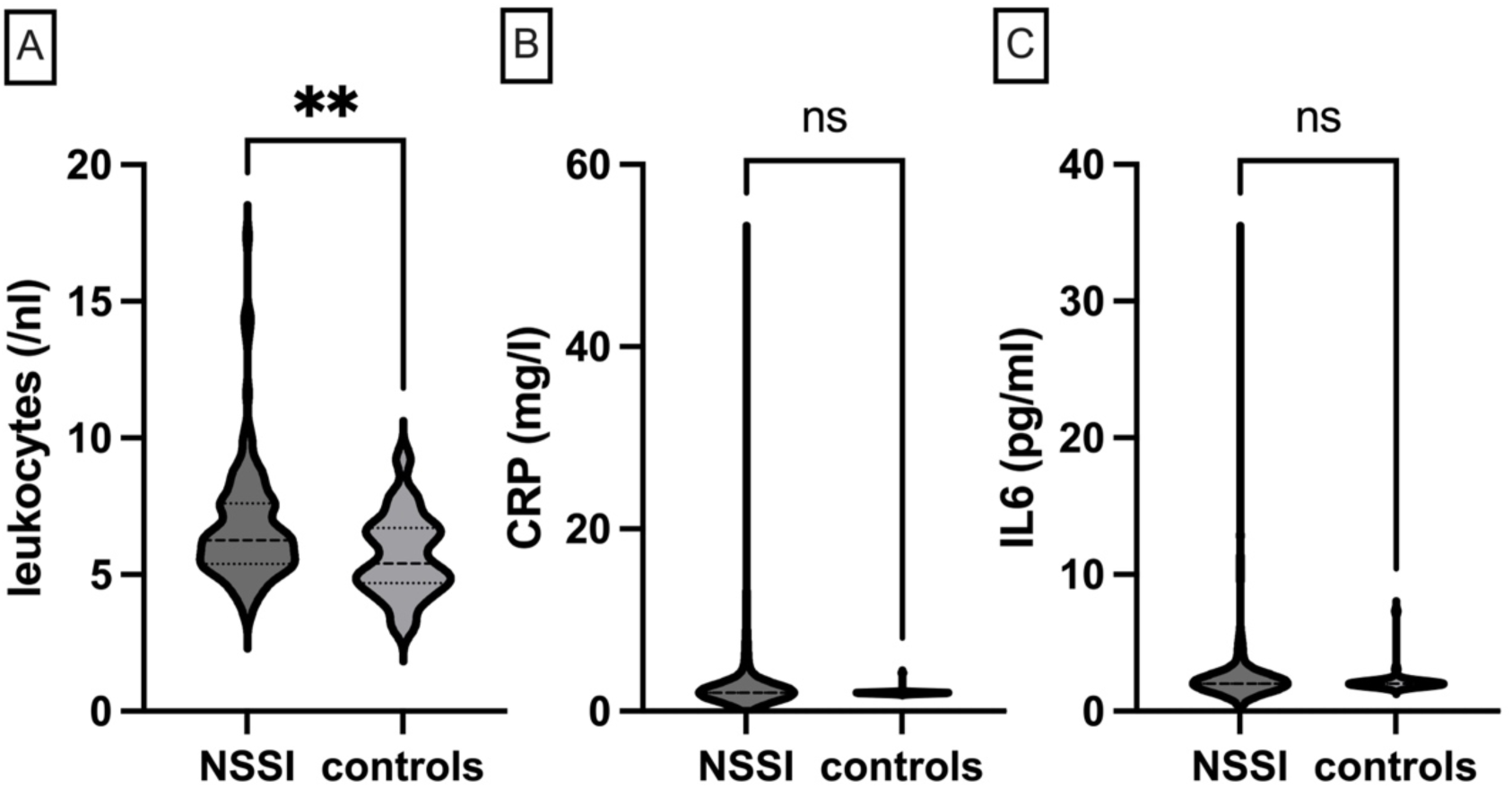
Inflammatory markers by group; CRP: c-reactive protein; IL6: interleukin-6; NSSI: non-suicidal self-injury; ns: non-significant

In the full sample, depression severity was negatively correlated with RMSSD (r = -.188, *p = .011*) (*Figure 3A*) and positively correlated with leukocyte (r = .219, *p = .005*) and CRP levels (r = .483, *p = .020*) (*Figures 3B and 3C*) when only observations above the detection threshold were included (CRP ≥ 2). However, IL-6 levels where only observations above the detection threshold were included (IL-6 ≥ 2) (r = .052, *p = .517*) and HR (r = .140, *p = .061*) were not associated with depression severity.

**Figure 3:**
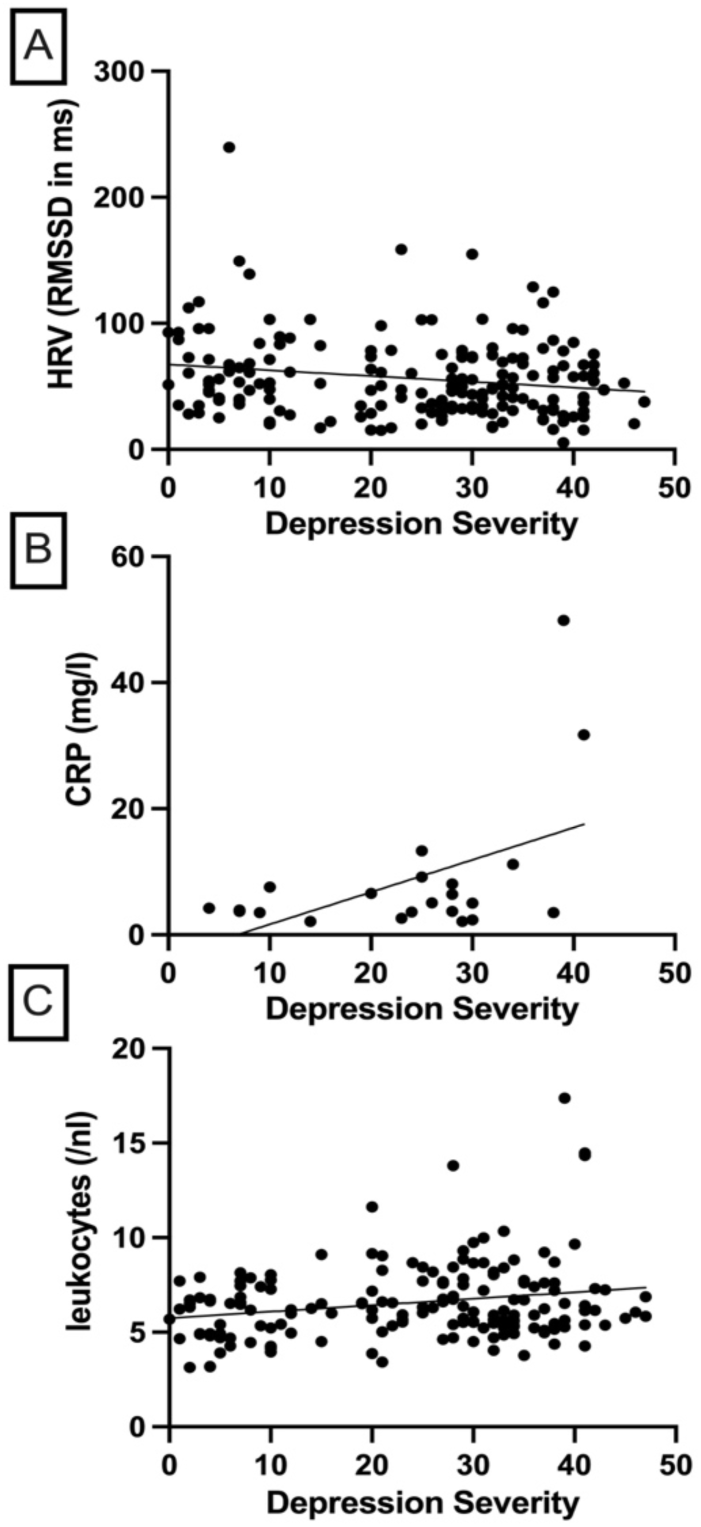
Association between Depression Severity, Heart Rate Variability and Inflammatory Markers; HRV: heart rate variability; RMSSD: root of the mean squared difference of successive inter-beat intervals in milliseconds; CRP: c-reactive protein

In the full sample, HRV was negatively correlated with leukocyte (r = -.159, *p = .031*) and CRP levels (r = -.456, *p = .022*) (*Figures 4A and 4B*) when only observations above the detection threshold were included (CRP ≥ 2). HR was positively correlated with leukocyte (r = .222, *p = .003*) and CRP levels (r = .637, *p = .001*) (*Figures 4C and 4D*) when only observations above the detection threshold were included (CRP ≥ 2). However, IL-6 levels were not related to neither HRV (r = -.151, *p = .395*) nor HR (r = -.016, *p = .928*).

**Figure 4:**
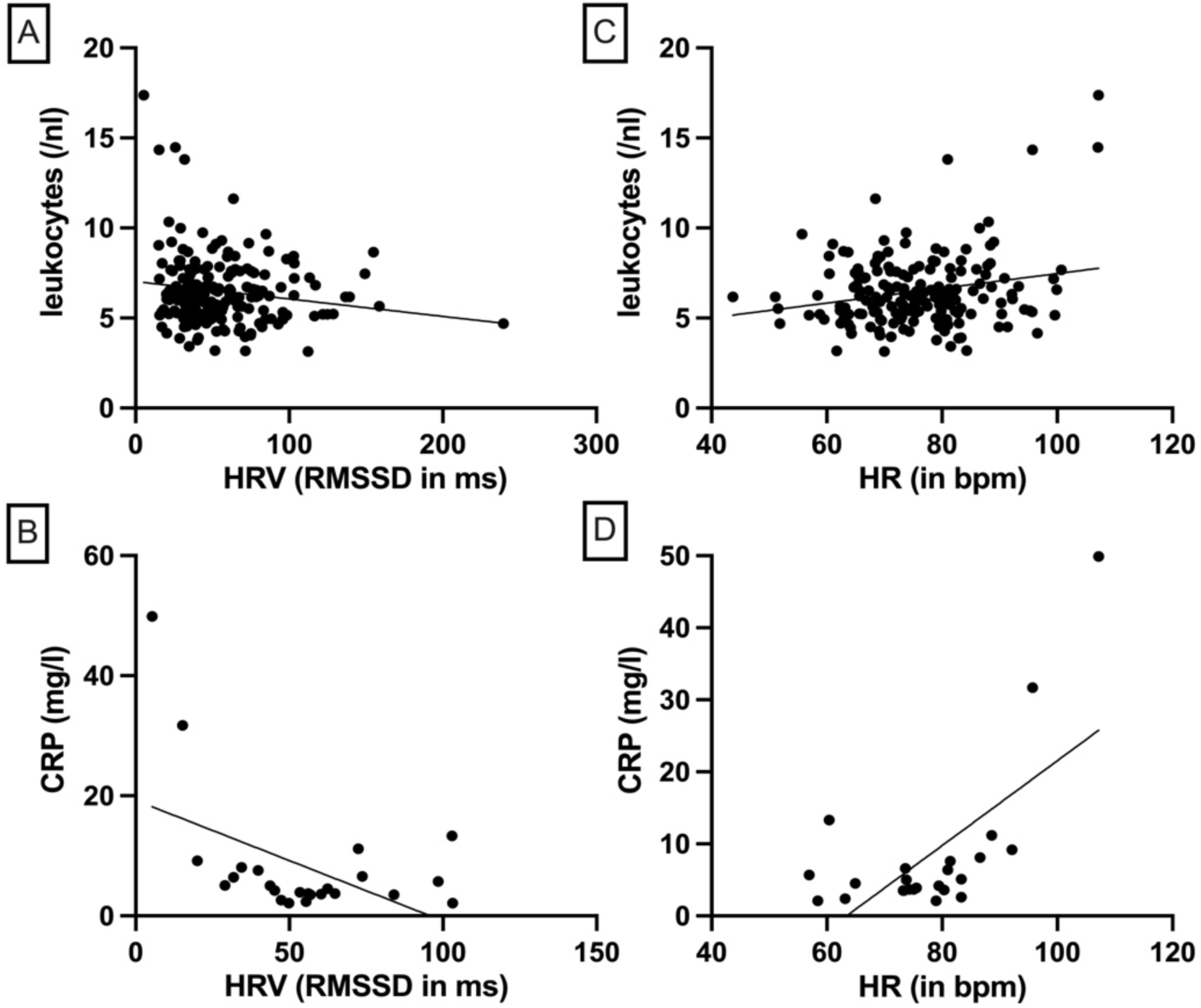
Association between Heart Rate and its Variability with Inflammatory Markers; HRV: heart rate variability; RMSSD: root of the mean squared difference of successive inter-beat intervals in milliseconds; HR: heart rate; CRP: c-reactive protein

Depression severity was not a significant mediator of the association between HRV and IL-6 (ab = -0.001, 95% CI [-0.003; 0.001]). Further, the direct effect of HRV on IL-6 was not significant (c′ = -0.005, 95% CI [− 0.021; 0.012]). Considering the association of HRV and CRP, depression severity was not a significant mediator (ab = -0.002, 95% CI [-0.008; 0.000], and the direct effect of HRV on CRP was also not significant (c′ = -0.018, 95% CI [-0.039; 0.003]). *Figure 5* depicts the mediation analysis with depression severity as a mediator of the associations between HRV and leukocytes. The analysis revealed that depression severity was a significant mediator of this association (ab = -0.002, 95% CI [-0.005; -0.000]). However, the direct effect of HRV on leukocytes was non-significant in this model (c′ = -0.008, 95% CI [- 0.017; 0.002]).

**Figure 5:**
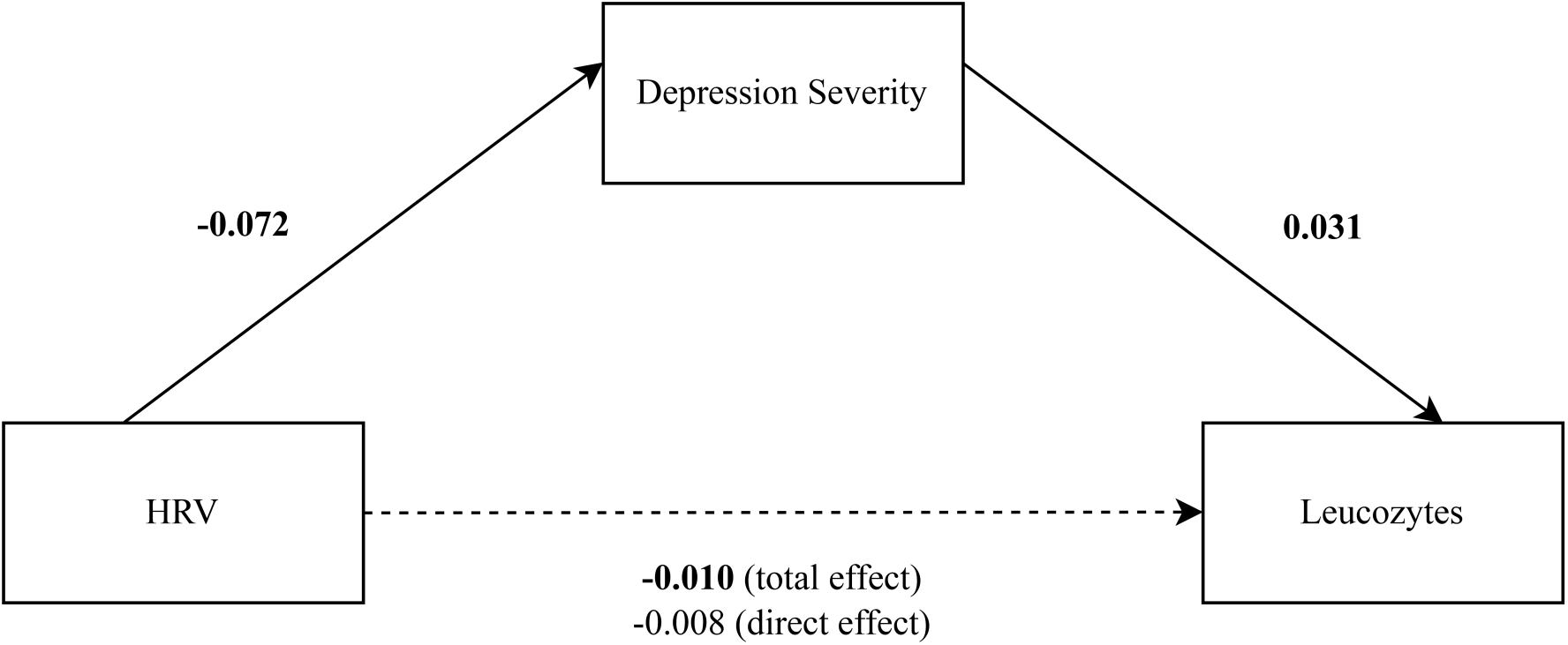
Mediation Model; HRV: heart rate variability (RMSSD in ms)

## Discussion

This study aimed to investigate the relationship between vagal activity, inflammatory markers and depression in adolescents engaging in NSSI compared to healthy controls to address whether reductions in vagal activity (indexed by HRV) were associated with increases in inflammatory markers, as suggested by the cholinergic anti-inflammatory pathway (Tracey, 2002).

In partial support of our first hypothesis, adolescents engaging in NSSI exhibited greater depressive symptoms, increased leukocyte counts and lower HRV compared to the control group. However, contrary to our hypothesis, IL-6 and CRP were not significantly elevated in the NSSI group.

Previous research has repeatedly demonstrated a positive association between depressive symptoms and NSSI (Hankin & Abela, 2011; Kindler et al., 2022), with one study suggesting that depressive symptoms may predict increases in NSSI (Marshall et al., 2013). It is assumed that depression and NSSI may co-occur as they share similar risk factors (e.g., genetic factors resulting in a reduced serotonin transmission (Klonsky, 2011) and cognitive factors such as negative attributional styles (Guerry & Prinstein, 2009; Marshall et al., 2013).

Our finding that leukocytes were significantly higher in the NSSI group is consistent with a prior report from the same study (Kindler et al., 2022), but in contrast to a recent study of hospitalized adolescent boys and girls (Zheng et al., 2022). Our findings which also demonstrate no group differences in CRP and IL-6 is consistent with prior research (Kim et al., 2020; Kindler et al., 2022). However, it is important to acknowledge the potential mediating or confounding effect of smoking which was significantly increased in the NSSI sample. Whilst significant group differences in smoking levels were controlled for in our analyses, it is important to note that smoking is associated with increased CRP, leukocyte count and IL-6 (Aldaham et al., 2015; Ohsawa et al., 2005; Sunyer et al., 1996) and that there may be a bidirectional relationship between depression and smoking in adolescents (Chaiton et al., 2009). Thus, our null findings for leukocyte and IL-6 warrant further investigation to better elucidate whether smoking mediates or confounds the relationship between depression and inflammation. The finding that HRV (as measured by RMSSD) was significantly lower in the NSSI group build on a previous study in which respiratory sinus arrhythmia was measured in a smaller sample of parasuicidal adolescent girls (Crowell et al., 2005), but contrast with another study that found only an inverse correlation with BPD symptomatology and not with NSSI (Koenig, Rinnewitz, et al., 2017). The latter study used a smaller sample in which the NSSI sample had reported more acts of NSSI in the past 12 months and fulfilled more BPD criteria than the sample in the current study (Koenig, Rinnewitz, et al., 2017).

In partial support of our second hypothesis, we found depression severity was positively correlated with leukocyte and CRP levels and negatively correlated with HRV. These findings align with data from previous research conducted in adolescent populations (Colasanto et al., 2020; Puangsri & Ninla-Aesong, 2021; Uçar et al., 2018). Contrary to our hypotheses, IL-6 was not related to depression severity. This is in contrast to results from a meta-analysis which found cross-sectional and bidirectional longitudinal associations between IL-6 and depression in adolescents (Colasanto et al., 2020). Despite the small number of studies in the subgroup analysis of longitudinal studies, we suspect our findings may be related to the role that IL-6 plays in the acute-phase response which may make it susceptible to short-term fluctuations caused by triggers such as infections (Latham et al., 2022). In partial support of our final hypothesis, we found depression severity mediated the associations between reduction in HRV and increases in leukocyte count. We also found a positive correlation between HRV and leukocytes as well as CRP. While IL-6 also showed no association with HRV, a recent meta-analysis found no significant association in subgroup analyses when RMSSD was used as a cardiac marker (Williams et al., 2019). Our study is the first to lend support to the ‘inflammatory reflex’ in an adolescent sample, that is, a dysregulated ANS is associated with raised inflammatory markers in depressed adolescents. This association is confirmed by a recent study with adults which found lower HRV (as indexed by respiratory sinus arrhythmia) was associated with an increased concentration in inflammatory cytokines (i.e., TNFa, IL6, Il1a and IFNg) (Buchmann et al., 2022). Future research should investigate these associations longitudinally to examine whether this relationship is maintained over time with improvement in depressive symptoms following psychiatric treatment. Further research should also examine whether adjuvant neurobiological treatments may modulate a dysregulated ANS and subsequently produce reductions in depression and inflammatory markers.

## Limitations

Our study has a few limitations that need to be considered. The cross-sectional design precludes any conclusions that may suggest a causal relationship. Further longitudinal research would be useful to clarify the directionality of the relationships between depression, HRV and inflammation in adolescents with NSSI. Studies addressing the impact of anti-inflammatory agents on these associations would allow such conclusions to be drawn (Vöckel et al., 2024). We were also unable to analyse all available peripheral inflammatory markers, some of which have been investigated in previous studies (Buchmann et al., 2022; Kim et al., 2020; Zheng et al., 2022). Further, the immunoassay used to determine inflammatory markers based on blood draws had a high detection threshold, precluding robust conclusions when it comes to CRP and IL-6 that were only measurable in a small subset of the sample. Finally, the generalizability of our findings to male adolescent is limited as only female adolescents were included in the study.

## Conclusion

In line with the ‘inflammatory reflex’ hypothesis, our study supports the notion that a dysregulated ANS may be associated with elevated inflammatory markers in a sample of adolescents with NSSI. We also found that adolescents with NSSI are more likely to show elevated leukocyte levels and lower HRV when compared to healthy controls and that this association is likely mediated by depression severity. Whilst further research should study these associations in a longitudinal manner with a greater array of high-sensitive inflammatory markers, our findings demonstrate the potential for the use of neurobiological and inflammatory biomarkers in the assessment of mental disorders in adolescents.

**Table 1:** Sample Characteristics by Group; school: Gymnasium refers to the highest level/track of secondary education, Realschule is the middle level/track while Hauptschule is the lowest level/track

|  | NSSI | controls | missings | *p* |
| --- | --- | --- | --- | --- |
| n | 154 | 46 |  |  |
| Age, m (SD) | 14.96 (1.51) | 14.71 (1.29) | 0/0 | .322 |
| BMI, m (SD) | 21.28 (3.75) | 20.27 (3.11) | 0/0 | .107 |
| Smoked today, n (%) | 18 (11.69) | 0 (0) | 0/0 | .015 |
| Current school, n (%)<sup>3</sup> |  |  | 0/0 | .001 |
| *Gymnasium* | 52 (33.77) | 29 (63.04) | 0/0 | - |
| *Hauptschule* | 56 (36.36) | 14 (30.43) | 0/0 | - |
| *Realschule* | 17 (11.03) | 2 (4.35) | 0/0 | - |
| *other* | 29 (18.83) | 1 (2.17) | 0/0 | - |
| BPD criteria, mean (SD) | 3.42 (2.09) | 0.07 (0.33) | 0/0 | .001 |
| Depression severity, mean (SD) | 30.16 (8.98) | 6.38 (4.87) | 18/1 | .001 |
| NSSI past 12 months, mean (SD) | 70.97 (73.84) |  | 0/0 |  |
| On medications, n (%) | 27 (17.53) | 2 (4.35) | 0/0 | .026* |
| CRP, mean (SD) | 3.04 (4.91) | 2.05 (0.33) | 10/2 | .185 |
| Leukocytes, mean (SD) | 6.71 (2.06) | 5.69 (1.50) | 14/10 | .003 |
| IL-6, mean (SD) | 2.76 (3.31) | 2.18 (0.84) | 22/5 | .271 |
| HRV, mean (SD) | 55.19 (29.15) | 66.22 (39.69) | 0/0 | .041 |
| HR, mean (SD) | 76.32 (10.50) | 72.89 (10.96) | 0/0 | .056 |

## Data Availability

All data produced in the present study are available upon reasonable request to the authors.

## Notes

### Competing Interest Statement

The authors have declared no competing interest.

### Funding Statement

This study was funded by the Dietmar Hopp Foundation.

### Author Declarations

The scientific evaluation of AtR!Sk was approved by Faculty of Medicine Ethics Committee at the University of Heidelberg (IRB approval number S-449/2013, and the additional neurobiological assessments: IRB approval number S-514/2015).

