## Supplementary Material for "The Cholinergic Anti-Inflammatory Reflex as a Function of Depression Severity in Adolescent Non-Suicidal Self-Injury"

**Supplementary Table 1. Mixed-effects multilevel regression, controlling for smoking and medication use in group differences**

| | Adjusted coefficient (95% confidence interval) | *p-value* |
|---|---|---|
| CRP | 1.13 (-0.34, 2.61) | *.133* |
| Leukocytes | 0.90 (0.23, 1.57) | *.009* |
| IL-6 | 0.49 (-0.53, 1.52) | *.346* |
| Depression severity | 23.62 (20.81, 26.43) | <*.001* |
| HRV | -11.80 (-22.48, -1.12) | *.030* |
| HR | 3.33 (-0.24, 6.89) | *.067* |
